# CoMix: comparing mixing patterns in the Belgian population during and after lockdown

**DOI:** 10.1101/2020.08.06.20169763

**Authors:** Pietro Coletti, James Wambua, Amy Gimma, Lander Willem, Sarah Vercruysse, Bieke Vanhoutte, Christopher I Jarvis, Kevin Van Zandvoort, John Edmunds, Philippe Beutels, Niel Hens, the SIMID-COVID19 Team

## Abstract

**Background:** The COVID-19 pandemic has shown how a newly emergent communicable disease can lay considerable burden on public health. To avoid system collapse, governments have resorted to several social distancing measures. In Belgium, this included a lockdown and a following period of phased re-opening.

**Methods:** A representative sample of Belgian adults was asked about their contact behaviour from mid-April to the beginning of August, during different stages of the intervention measures in Belgium. Use of personal protection equipment (face masks) and compliance to hygienic measures was also reported. We estimated the expected reproduction number computing the ratio of R_0_ with respect to pre-pandemic data.

**Findings:** During the first two waves (the first month) of the survey, the reduction in the average number of contacts was around 80% and was quite consistent across all age-classes. The average number of contacts increased over time, particularly for the younger age classes, still remaining significantly lower than pre-pandemic values. From the end of May to the end of July, the estimated reproduction number has a median value larger than one, although with a wide dispersion. Estimated _0_ fell below one again at the beginning of August.

**Conclusions:** We have shown how a rapidly deployed survey can measure compliance to social distancing and assess its impact on COVID-19 spread. Monitoring the effectiveness of social distancing recommendations is of paramount importance to avoid further waves of COVID-19.

## 1 Introduction

The COVID-19 pandemic due to the novel coronavirus (SARS-CoV-2) has shown how newly emerging infectious diseases can lay considerable burden on public health and social economic welfare of the society. Since its emergence, over 40 million confirmed cases and 1,250,000 deaths have been recorded as of October 21st 2020 [1]. In the absence of established pharmaceutical interventions, many countries across the globe have resorted to non-pharmaceutical interventions, advocacy of proper hygienic measures (hand washing, sanitizing), as well as promotion of wide-spread usage of masks to help combat the spread of this disease. However, sustainability of some of the imposed measures is infeasible in the long term, due to an urgent need to returning back to normal social life as well as rekindling the economy. Thus governments have been prompted to lift some of the measures in a phased manner whilst enforcing new/existing rules such as wearing masks in designated places such as in public transport, hospitals, schools, workplaces and other places that attract large crowds and gatherings.

As COVID-19 is primarily transmitted through close-contact interaction with infected individuals [2], data on social contacts is indispensable in informing mathematical modeling studies being employed to explore the evolution of this disease. The last decade of research in infectious disease modeling has shown how quantifying contact patterns is crucial to capture disease dynamics [3]. However, social contact data capturing behavioral changes in the population during and across different stages of an epidemic is mostly lacking and mathematical models need to rely on various assumptions, which might be unverifiable. This raises validity concerns on their appropriateness in guiding decision making.

Thus, as many governments are carefully monitoring the situation to avoid further waves of COVID-19, continual data collection is vitally important to closely monitor changes in social mixing. This can provide insights on the impact of different intervention measures as well as help in real-time management of the COVID-19 crisis, together with other insights from social and behavioral sciences [4].

Studies comparing social contact patterns before and during the COVID-19 pandemic have been reported for Wuhan and Shanghai [5], the UK [6], the Netherlands [7], Luxembourg [8], the US [9] and in multiple countries (Belgium, France, Germany, Italy, the Netherlands, Spain, the UK, and the US) [10]. The overall reduction in the total number of contacts made by individuals ranged from 48% to 85%, stressing once again the importance of quantifying the impact of social distancing separately for each country. Also, although little variations in the number of contacts over time were measured [10] up to mid-April, this may change as countries relieve stricter measures and social interactions need to adjust to the new post-lockdown reality [8].

In this paper, we present results from a longitudinal survey of the adult population in Belgium, representative by age, gender and region of residence. The survey involves multiple waves of data collection, and is part of a wider study to look at changes in contact patterns across European countries (see e.g. UK [6]). Here, we present results for eight waves (= 16 weeks). We quantify the changes in social contact patterns comparing pre-pandemic, lockdown and post-lockdown periods and its impact on the transmission dynamics of COVID-19 based on the changes in the basic reproduction number relying on the next generation principle. We use a published survey of the Flemish region (Belgium) conducted in 2010 [11, 12] as reference for the pre-pandemic social mixing. Also, we assess the uptake of face mask wearing and adherence to hygienic measures in the population over time.

## 2 Methods

### Ethics statement

Participants (aged 18 years or older) opt-in for the study voluntarily. Informed consent was collected and all analyses were performed on pseudo-anonymised data. The study was approved by the ethics committee of the University of Antwerp (reference number EC UZA 20/13/147).

### Survey methodology

A representative sample of Belgian adults was asked to participate in a multiwave survey about their contact behavior. The sample was selected from existing panels of individuals frequently participating in online surveys and selected to be representative of the Belgian population using quotas on age, gender and region of residence (see Supporting Information for more details). Here we present results for the first eight waves that were collected during and after the lockdown, with different intervention strategies taking place (see Figure 1 for details). Participants’ age, education level and occupation were recorded, together with information regarding their socio-economic status, health status, whether they experienced symptoms, attitude towards intervention measures and adherence to intervention measures. Age, gender, occupation and symptoms experienced were collected also for household members. Specific questions on whether the participants or any of their household members tested positive for COVID-19 or if they knew somebody who did were asked. In addition, self-isolation and quarantine (i.e. isolation after potential exposure to infected individuals) periods in the last 7 weeks were reported. During the survey day, participants were asked to report all contacts made between 5 am the day preceding the survey and 5 am of the day of the survey. A contact was defined as an in-person conversation of at least a few words, or a skin-to-skin contact. Participants could report individual contacts or (from wave 3 onward) contacts with a group of individuals. For every individual contact, participants filled in the age and the gender of the contacted person, whether the contact included skin-to-skin touching, the duration of the contact and the frequency with which they usually contact this person. Information on the location was collected using pre-specified locations (home, work, school, leisure activities, other places) and specifying whether the contact took place in open air or indoors. Contacts reported in group could be marked as taking place at work, school or any other location and allowed only for three age categories (0-18, 19-65 and 65+ years) and were reported in two categories: one explicitly stating that they involved physical contact and one without specifying this. We assigned specific age-values to the contacts in these two categories by sampling from the age-distribution of the reported individual contacts. Contact duration and frequency were not collected for contacts reported in groups.

**Figure 1:**
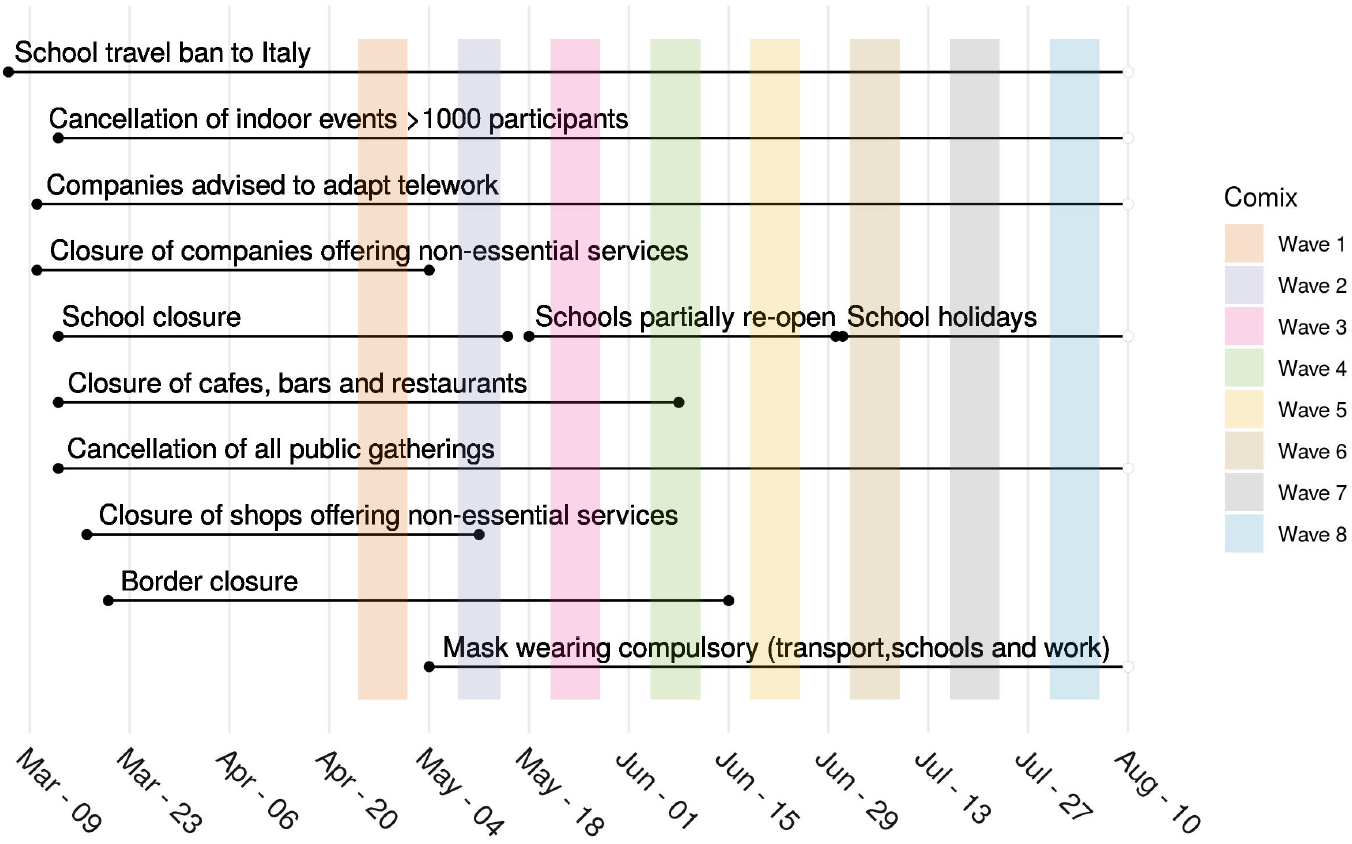
Calendar of interventions and CoMix wave data collections.

Participants could complete the survey using a website or an app on their phone/tablet.

## Data analysis

We computed the average number of contacts per participant, stratified by several participant characteristics, like age, gender, household size and day of the week. We present results in comparison to a social contact survey conducted in the Flemish region of Belgium in 2010 [11, 12].

We computed R_0_ as the leading eigenvalue of the next generation matrix informed by the contact matrix while assuming the transmission rates to be proportional to the social contact rates, i.e. “the social contact hypothesis” [13, 14] from the 2010 survey and from the eight waves of the CoMix survey separately. To account for sampling variability, we generated 10,000 contact matrices via bootstrapping using the SocialMixr package [15]. Then, we computed the R_0_ ratio with respect to the pre-pandemic period as the ratio between R_0_ from CoMix survey and R_0_ from the 2010 survey for each bootstrap iteration, ending up with a distribution of R_0_ ratios [16]. We computed the ratio using different sets of contacts: the total number of contacts and only contacts involving skin-to-skin touching.

As the survey does not include individuals younger than 18 years of age, we also assessed the impact of imputing social contacts for the 0-4 and 5-17 age classes on the ratio of R_0_. To impute these contacts we followed the approach described in [17] and applied to another social contact survey collected within the same initiative [6]. We first computed the R_0_ ratio comparing the CoMix survey with the 2010 survey for each contact location, considering only the age classes in common. Then, we computed the number of contacts made by individuals in the 0-4 and 5-17 age classes for the CoMix survey as the corresponding number of contacts in the 2010 survey, multiplied by the eigenvalue ratio in each contact location. We again used bootstrap (n=10,000) to account for sampling variability. These *imputed* contact matrices are then used to compute the R_0_ ratio as explained before. We compare the results of both approaches. R version 3.6.3 was used for all analyses. The code used in this analysis is based on SocialMixr package [15] and the Socrates tool [18, 19]. The social contact data is available on Zenodo [20].

## 3 Results

### 3.1 Sample composition

During the first wave, 1542 participants took part in the survey, divided among 732 males (47.5%) and 810 females (52.5%) (Table 1). Table S2 presents information on participation rates for each wave. The average participant’s age was 48.4 years (standard deviation (sd) =16.3 years), with a median age of 50 years, and an inter-quartile range (IQR) of [35-65]. The average household size was 2.8 (sd = 1.4), IQR [2-4] with a maximum household size of 10. In total, data on 4290 household members, including the participants, was collected.

**Table 1:** Summary of sample characteristics.

|  | Population<br>(1 Jan 2020) | Wave 1<br>(N=1542) | Wave 2<br>(N=1277) | Wave 3<br>(N=1142) | Wave 4<br>(N=951) | Wave 5<br>(N=924) | Wave 6<br>(N=902) | Wave 7<br>(N=760) | Wave 8<br>(N=833) |
| --- | --- | --- | --- | --- | --- | --- | --- | --- | --- |
| Region |  |  |  |  |  |  |  |  |  |
| Brussels region | 10.6% | 110 (7.1%) | 81 (6.3%) | 75 (6.6%) | 60 (6.3%) | 57 (6.2%) | 56 (6.2%) | 48 (6.4%) | 52 (6.3%) |
| Flanders | 57.7% | 961 (62.3%) | 825 (64.6%) | 732 (64.1%) | 590 (62.0%) | 572 (61.9%) | 561 (62.2%) | 496 (65.2%) | 525 (63.0%) |
| Wallonia | 31.8% | 471 (30.5%) | 371 (29.1%) | 335 (29.3%) | 301 (31.7%) | 295 (31.9%) | 285 (31.6%) | 216 (28.4%) | 256 (30.7%) |
| Age group |  |  |  |  |  |  |  |  |  |
| 0-9 | 11.1% | - | - | - | - | - | - | - | - |
| 10-19 | 11.3% | 35 (2.3%) | 20 (1.6%) | 11 (0.96%) | 8 (0.84%) | 10 (1.1%) | 8 (0.89%) | 4 (0.5%) | 4 (0.5%) |
| 20-29 | 12.3% | 229 (14.9%) | 156 (12.2%) | 123 (10.8%) | 89 (9.4%) | 95 (10.3%) | 84 (9.3%) | 66 (8.7%) | 78 (9.4%) |
| 30-39 | 13.0% | 222 (14.4%) | 183 (14.3%) | 157 (13.7%) | 124 (13.0%) | 121 (13.1%) | 124 (13.7%) | 82 (10.8%) | 102 (12.2%) |
| 40-49 | 13.1% | 280 (18.2%) | 233 (18.2%) | 209 (18.3%) | 156 (16.4%) | 152 (16.5%) | 145 (16.1%) | 129 (17.0%) | 138 (16.6%) |
| 50-59 | 13.8% | 310 (20.1%) | 273 (21.4%) | 253 (22.2%) | 221 (23.2%) | 198 (21.4%) | 200 (22.2%) | 177 (23.3%) | 195 (23.4%) |
| 60-69 | 11.7% | 318 (20.6%) | 286 (22.4%) | 271 (23.7%) | 243 (25.6%) | 237 (25.6%) | 231 (25.6%) | 202 (26.6%) | 207 (24.8%) |
| 70+ | 13.7% | 148 (9.6%) | 126 (9.9%) | 118 (10.3%) | 110 (11.6%) | 111 (12.0%) | 110 (12.2%) | 100 (13.1%) | 109 (13.1%) |
| Gender |  |  |  |  |  |  |  |  |  |
| Males | 49.3% | 732 (47.5%) | 604 (47.3%) | 560 (49.0%) | 484 (50.9%) | 463 (50.1%) | 445 (49.3%) | 386 (50.8%) | 429 (51.5%) |
| Females | 50.7% | 810 (52.5%) | 671 (52.5%) | 581 (50.9%) | 467 (49.1%) | 460 (49.8%) | 456 (50.6%) | 374 (49.2%) | 399 (47.9%) |
| NAs | - | - | 2 (0.16%) | 1 (0.09%) | - | 1 (0.11%) | 1 (0.11%) | - | 5 (0.6%) |

Nearly half of the participants were living with children (51.8%, 799 participants), 22.5% (347 participants) were part of a childless couple and 16.0% (247 participants) were living alone. Few participants (1.0%, 16 participants) reported living in a household with two or more families, 2.7% (41 participants) lived with two or more non-family adults and 6.1% (92 participants) reported living in yet a different type of household.

Overall, 34.4% (530) of the participants reported being fully employed, 9.3% (143) working part-time, 4.5% (70) being self-employed, 25.7% (396) being retired, 7.6% (117) being students and 5.3% (82) being unemployed. Additionally, 4.9% (75) of participants reported being homemakers and 8.4% (129) being disabled or with long term sickness.

In the following waves the participant sample varied in size, with minimal impact on the composition of the sample. The number of survey participants declined from of 1277 participants (82.8% of the total) during the second wave to 833 participants (54%) during the eighth wave.

### 3.2 Participant contacts

The average number of contacts in all the waves of the CoMix survey shows a marked reduction compared with the 2010 pre-pandemic data and also shows less disassortative mixing with age (Figure 2 and Table 2). During lockdown (wave 1), the reduction in the average number of contacts was as high as 80% with respect to the 2010 data. During wave 2 the number of contacts was in line with the lockdown results, although wave 2 was collected after the re-opening of companies and shops offering non-essential goods. From wave 3 to wave 7, a marked increase in the average number of contacts was observed, especially for the younger age classes. The average number of contact decreased again from wave 7 to wave 8. The median number of contacts however remained quite stable (see Figure SI 1), suggesting that the increase in the average number of contacts is due to a limited number of individuals that report a large number of contacts. Participants living in the Flemish region reported a higher number of contacts with respect to participants in the Brussels and Walloon regions (see Figure SI 5). Over time, we also observed a reduction in the average number of contacts made at home, whereas the number of contacts made away from home increased (Figure 3). This is consistent with contacts being established for the vast majority at home during lockdown. When lockdown was relieved, an increase of the percentage of contacts outside home was therefore expected. Contacts at home were also more likely to involve physical contact and to last longer (see Figure SI 6 and Figure SI 7) than contacts outside the household. With respect to the pre-pandemic period, in all CoMix survey waves a smaller fraction of contacts involving skin-to-skin touching was observed.

**Table 2:** Average number of contacts.

| Category | Value | CoMix survey |  |  |  |  |  |  |  | 2010 survey |
| --- | --- | --- | --- | --- | --- | --- | --- | --- | --- | --- |
|  |  | Wave 1<br>mean (IQR) | Wave 2<br>mean (IQR) | Wave 3<br>mean (IQR) | Wave 4<br>mean (IQR) | Wave 5<br>mean (IQR) | Wave 6<br>mean (IQR) | Wave 7<br>mean (IQR) | Wave 8<br>mean (IQR) |  |
| Age | Overall | 2.68 (1-4) | 2.87 (1-4) | 4.41 (1-4) | 4.89 (1-4) | 4.76 (1-4) | 5.76 (1-4) | 4.71 (1-5) | 3.48(1-4) | 18.41 (6-21) |
|  | 18-29 | 3.34 (1-4) | 3.38 (1-4) | 6.25 (1-5) | 6.55 (1-6) | 8.88 (1-6) | 7.61 (1-6) | 8.57(1-7) | 4.70(1-4) | 15.01 (8-19) |
|  | 30-39 | 3.14 (2-4) | 3.20 (1-4) | 5.35 (1-5) | 4.63 (1-4) | 4.12 (1-4) | 5.57 (1-6) | 5.71(0-5) | 2.86(0-4) | 13.84 (7-17) |
|  | 40-49 | 3.14 (1-4) | 3.39 (1-4) | 4.87 (1-5) | 6.85 (1-6) | 6.12 (1-5) | 6.92 (1-6) | 7.01(1-5) | 4.59(1-5) | 13.43 (7-17) |
|  | 50-59 | 2.45 (1-3) | 2.73 (1-4) | 3.91 (1-4) | 6.26 (1-6) | 4.96 (1-5) | 4.58 (1-5) | 4.13(1-4) | 3.59(1-4) | 12.66 (6-16) |
|  | 60-69 | 2.07 (1-3) | 2.51 (1-4) | 3.52 (1-4) | 2.96 (1-4) | 2.96 (1-4) | 4.01 (1-4) | 3.06(1-4) | 2.85(1-4%) | 10.15 (4-12) |
|  | 70+ | 1.70 (1-2) | 1.92 (1-3) | 3.27 (1-4) | 2.47 (1-3) | 3.17 (1-4) | 4.22 (1-5) | 2.61(1-3) | 2.71(1-4%) | 8.57 (3-12) |
| Gender | Female | 2.68 (1-4) | 3.08 (1-4) | 4.22 (1-4) | 4.7 (1-5) | 4.44 (1-5) | 5.32 (1-5) | 5.29 (1-5) | 3.18 (1-4) | 13.45 (6-18) |
|  | Male | 2.67 (1-4) | 2.64 (1-4) | 4.57 (1-4) | 5.08(1-4) | 5.09 (1-4) | 4.98 (1-5) | 4.16 (1-4) | 3.77 (1-4) | 13.62 (6-17) |
| Household size | 1 | 1.46 (0-2) | 1.55 (0-2) | 4.54 (0-2) | 3.35 (0-3) | 3.47 (0-3) | 2.93 (0-3) | 3.41 (0-2) | 1.86 (0-2) | 9.83 (4-13) |
|  | 2 | 1.92 (1-2) | 2.31 (1-3) | 3.42 (1-3) | 3.93 (1-4) | 4.13 (1-4) | 5.6 (1-5) | 4.52 (1-5) | 3.82 (1-4) | 11.54 (5-14) |
|  | 3 | 2.76 (2-3) | 2.90 (2-4) | 4.48 (2-4) | 6.51 (2-5) | 5.76 (2-4) | 5.63 (2-5) | 5.12 (2-5) | 3.51 (2-4) | 13.35 (6-17) |
|  | 4 | 3.57 (3-4) | 3.89 (3-5) | 5.53 (3-5) | 6.21 (3-6) | 5.74 (3-6) | 6.47 (3-6) | 6.11 (2-6) | 3.88 (2-4X) | 15.82 (8-20) |
|  | 5 | 4.81 (4-6) | 5.05 (4-6) | 5.95 (4-7) | 5.93 (4-6) | 5.61 (3-7) | 6.13 (2-7) | 6.46 (2-6) | 4.58 (1-6) | 17.12 (8-24) |
|  | 6 | 5.66 (5-5) | 6.77 (5-8) | 6.65 (4-7) | 13.69 (5-8) | 9.75 (5-9) | 8.94 (4-11) | 5.67 (5-9) | 11.1 (5-9) | 17.33 (9-22) |
|  | 7+ | 7.37 (6-8) | 6.41 (5-8) | 14.25 (5-13) | 6.5 (6-7) | 24.75 (5-28) | 4.67 (4-7) | 19.67 (12-29) | 7 (7-7) | 20.00 (9-22) |
| Weekday | Monday | 2.34 (1-3) | 3.69 (1-4) | 4.88 (2-6) | 4.99 (1-5) | 5.29 (2-4) | 6.21 (1-5) | 3.76(1-5) | 2.72(1-3) | 12.62 (6-16) |
|  | Tuesday | 3.51 (1-4) | 3.14 (2-5) | 7.19 (2-8) | 6.63 (1-5) | 5.64 (1-5) | 4.99 (1-5) | 7.44(1-7) | 3.40(1-5) | 14.63 (7-19) |
|  | Wednesday | 2.77 (1-4) | 3.03 (1-5) | 4.08 (1-4) | 4.59 (1-4) | 3.76 (1-4) | 4.09 (1-4) | 12.62(2-8) | 3.82(1-4) | 14.37 (7-20) |
|  | Thursday | 3.05 (1-4) | 3.26 (1-4) | 3.68 (1-4) | 4.22 (1-5) | 6.61 (1-5) | 11.22 (2-9) | 5.28(1-5) | 3.77(1-4) | 12.61 (6-16) |
|  | Friday | 3.21 (1-4) | 3.22 (2-4) | 4.53 (2-5) | 3.39 (1-5) | 3.54 (1-4) | 6.33 (1-4) | 3.06(1-4) | 2.36(1-3) | 14.77 (7-19) |
|  | Saturday | 2.49 (1-4) | 2.55 (1-4) | 5.29 (1-5) | 4.16 (1-4) | 5.12 (1-6) | 5.66 (1-6) | 2.59(1-3) | 2.68(1-4) | 14.42 (7-19) |
|  | Sunday | 2.56 (1-3) | 2.94 (1-4) | 5.94 (1-6) | 8.93 (1-7) | 6.44 (2-10) | 5.8 (1-6) | 6.68(1-3) | 4.67(1-4) | 10.52 (4-13) |

**Figure 2:**
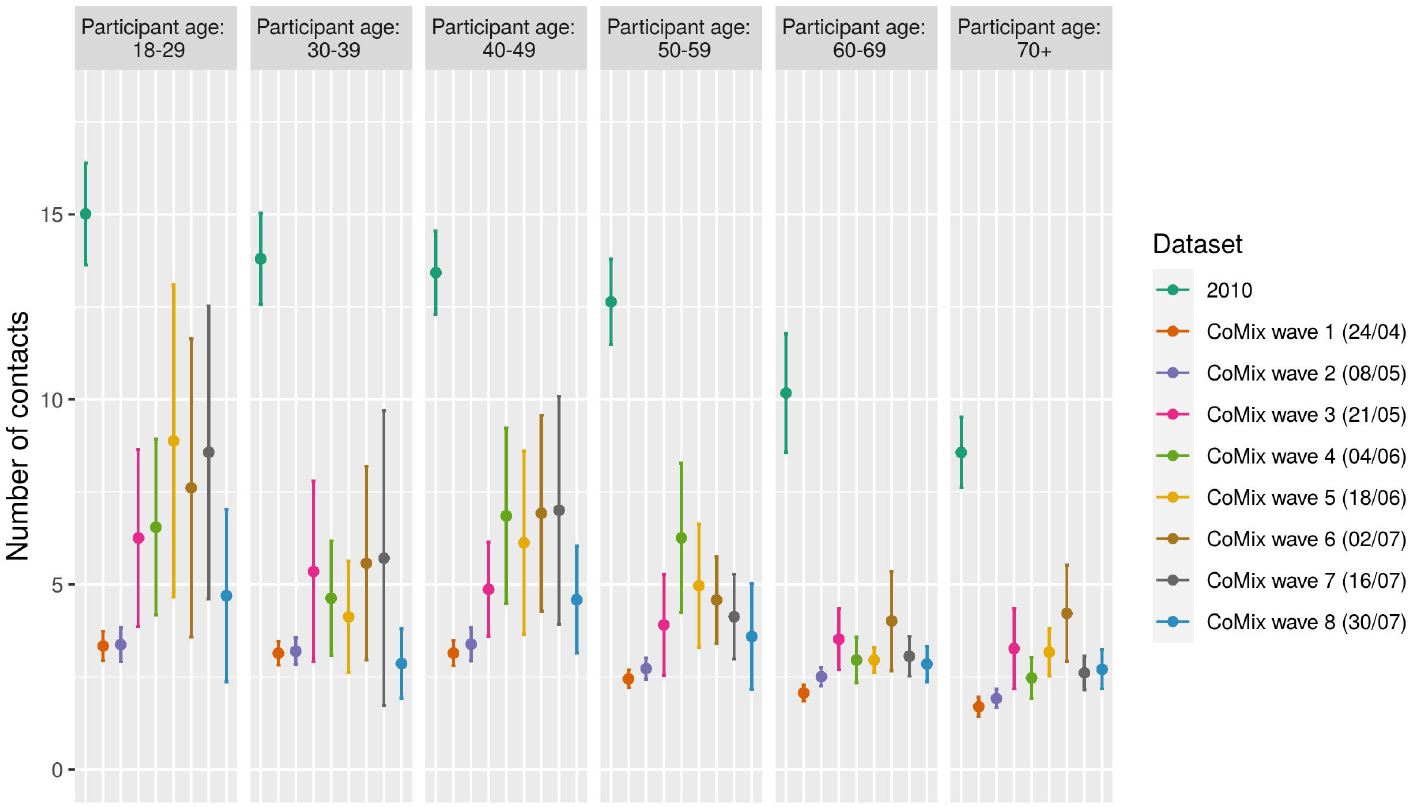
Contacts according to participant age class and dataset. Mean number of contacts broken down by participant age and dataset. For dataset coming from the CoMix survey, the starting date of the data collection is shown in brackets. Errorbars mark the 95% CI.

**Figure 3:**
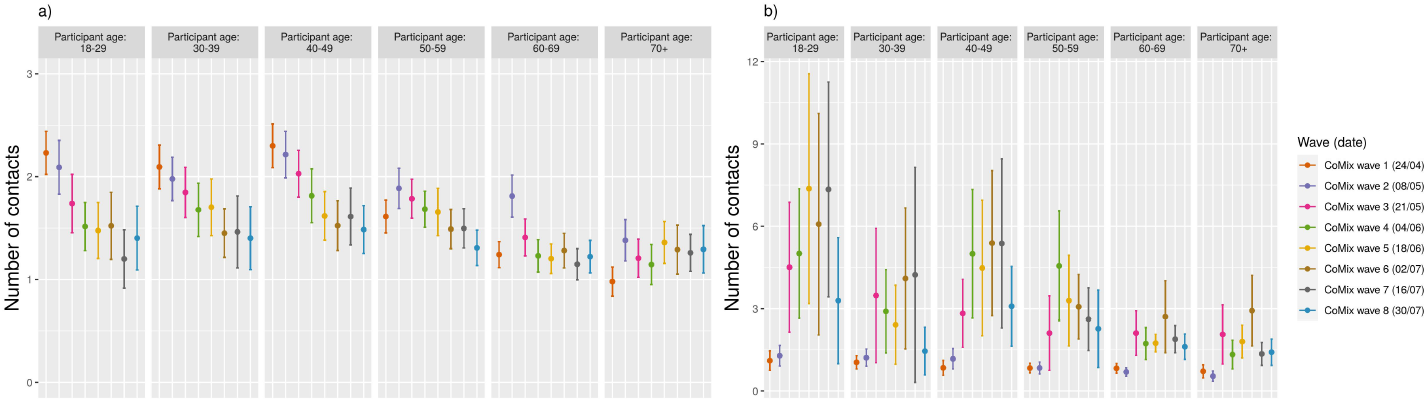
Home/away from home contacts. **(a):** Mean number of contacts at home. Errorbars mark the 95% CI.**(b):** Mean number of contacts away from home. Errorbars mark the 95% CI.

Participant gender, household size and day of the week do not show a significant relation with the average number of contacts. Figure 4 shows the contact matrices for the 2010 survey and the waves of the CoMix survey. The overall reduction in the number of contacts with respect to pre-pandemic scenario is marked by the color palette. Contact matrices from the CoMix survey present a more even structure, as they do not show a relatively higher contact intensity on the main diagonal of the matrix nor do they reveal work contacts (ranging from 18 years of age to 60 years of age) as the 2010 contact matrix does. From wave 3 onward, a larger number of contacts in the working population can be noticed, though with a weaker magnitude than during the pre-pandemic period. The high number of contacts established by participants in the youngest age category (from 18 years of age to 30 years of age) is clearly visible.

**Figure 4:**
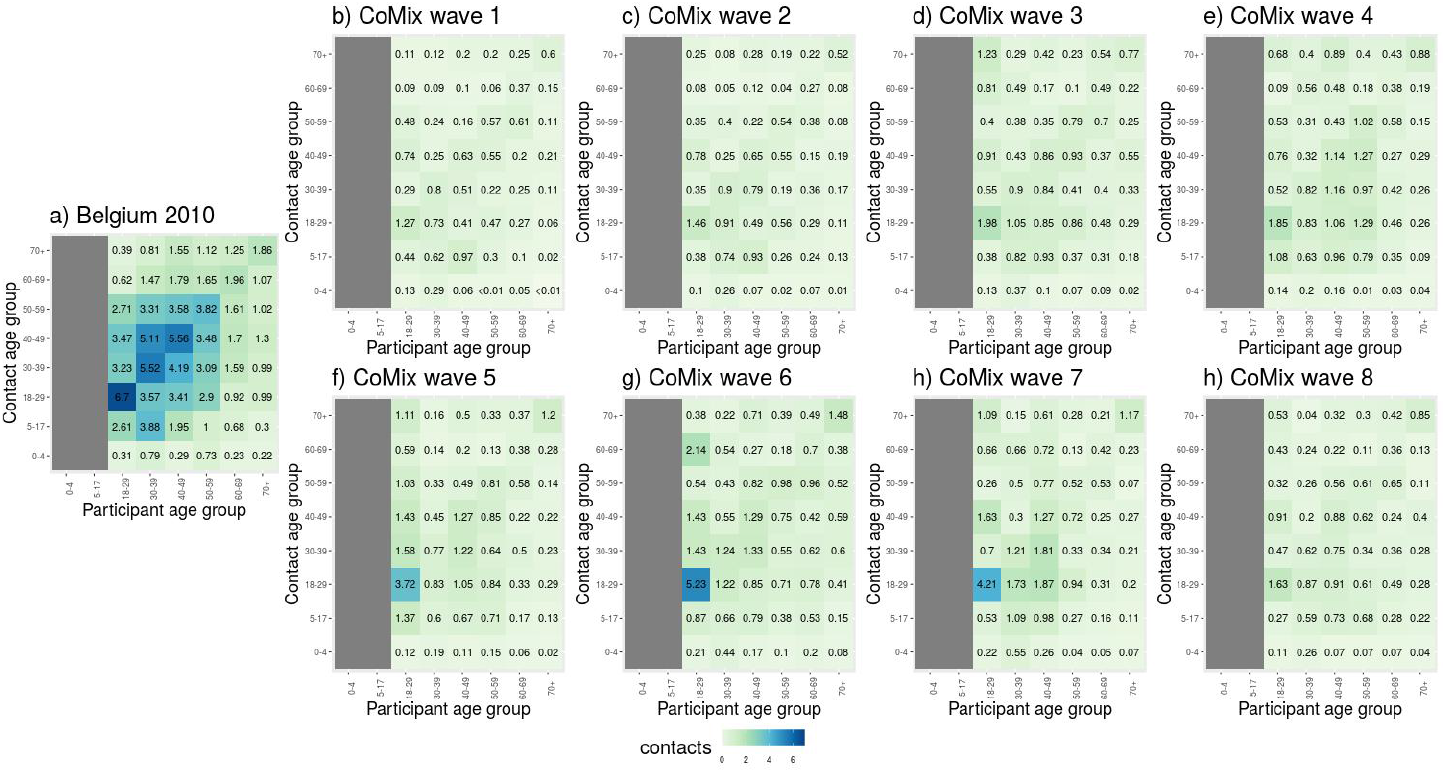
Social contact matrices. **(a):** Average number of daily reported contacts for the 2010 survey. **(b-g)** Average number of daily reported contacts for the 8 waves of the CoMix survey. The columns corresponding to participants below 18 years of age in the 2010 data have been removed, to ease the comparison with the CoMix data.

### 3.3 *R*_0_ reduction

We estimated R_0_ ratio between the eight waves of the CoMix survey and the 2010 data. The ratio increases with time up to mid July, ranging from a median of 0.186 (95% CI [0.171 : 0.203]) for wave 1 to a median of 0.474 (95% CI [0.344 : 0.644]) for wave 7 (Figure SI 13). At the end of July (wave 8) the R_0_ ratio decreases to 0.280 (95% CI [0.210 : 0.377]). When imputing contacts for children (i.e. individuals *<*18y), these numbers change to a median of 0.238 (95% CI [0.225 : 0.252]) for wave 1, 0.469 (95% CI [0.382 : 0.574]) for wave 7 and 0.308 (95% CI [0.269 : 0.353]) for wave 8 (Figure SI 13). Contact imputation generally increases the R_0_ ratio. Therefore, this means that contact imputation leads to a higher reproduction number during and after the lockdown. Considering the R_0_ to be normally distributed with mean 3.344 and standard deviation 0.028 [21], social contact data for wave 3 onward point towards a reproduction number that is higher than one both with and without contact imputation (Figure 5) up to mid July (wave 7). Data collected at the end of July (wave 8), instead, point towards a smaller reproduction number. When considering only physical contacts this increasing trend in the reproduction number is not present. It has to be considered, however, that the physical nature of the contact is not recorded for a large fraction of contacts from wave 3 onward (see Figure SI 3).

**Figure 5:**
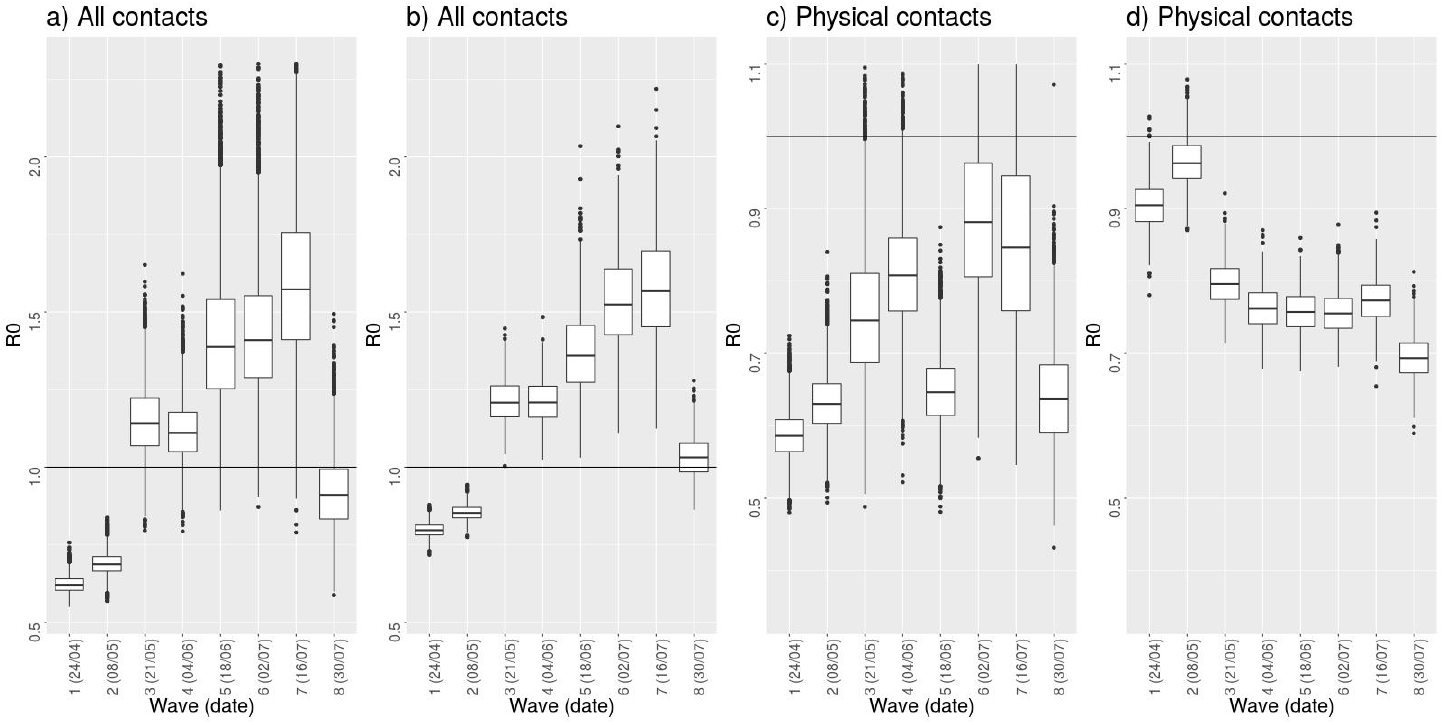
Reproduction number. **(a-b):** Reproduction number considering all contacts, without imputation (a) and with imputation of contacts for children (i.e. individuals *<*18y) (b).**(c-d):** Reproduction number considering physical contacts, without imputation (c) and with imputation of contacts for children (d). All values are obtained starting from a normally distributed R_0_ (mean= 3.344, sd=0.028) and the ratio of R_0_ computed with respect to the 2010 survey data. The horizontal line marks a reproduction number of 1.

### 3.4 Face mask wearing and hand hygiene

The fraction of participants reporting to have used a mask in the previous day in any one or more of the following; everywhere, walking on the street, cycling, during public transport, on the supermarkets/shops, during cinema/bar/restaurant, while at home, or any other place increased strongly from the first wave up to wave eight. Whereas during wave one only 18% of all participants reported wearing a mask, more than 75% reported wearing a mask during wave eight. Small variations are observable among different age classes (Figure 6), however non significant. On the other hand, the number of participants that reported washing their hands at least once in the last three hours slightly decreased over time, dropping from 92% during wave 1 to 89% during wave eight. In this case, significant differences are observed among age classes, in particular with younger individuals being less likely to wash their hands, consistently over all waves. No significant impact of gender is found on face mask wearing, nor on hands washing (Figure SI 14 and SI 15).

**Figure 6:**
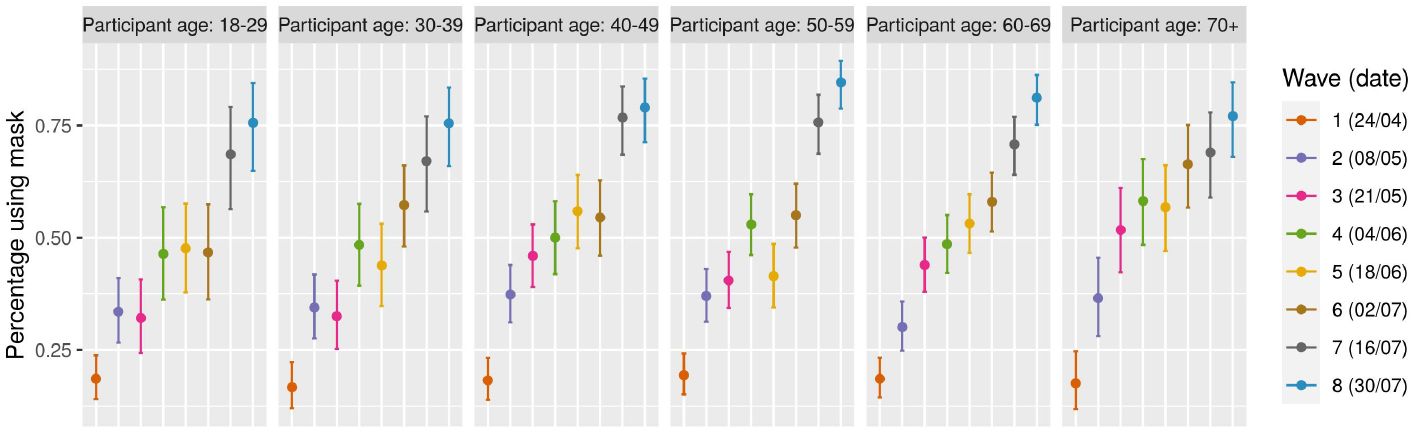
Percentage of participants using face mask. Average number of participants wearing mask broken down by age. Errorbars mark the 95% CI.

**Figure 7:**
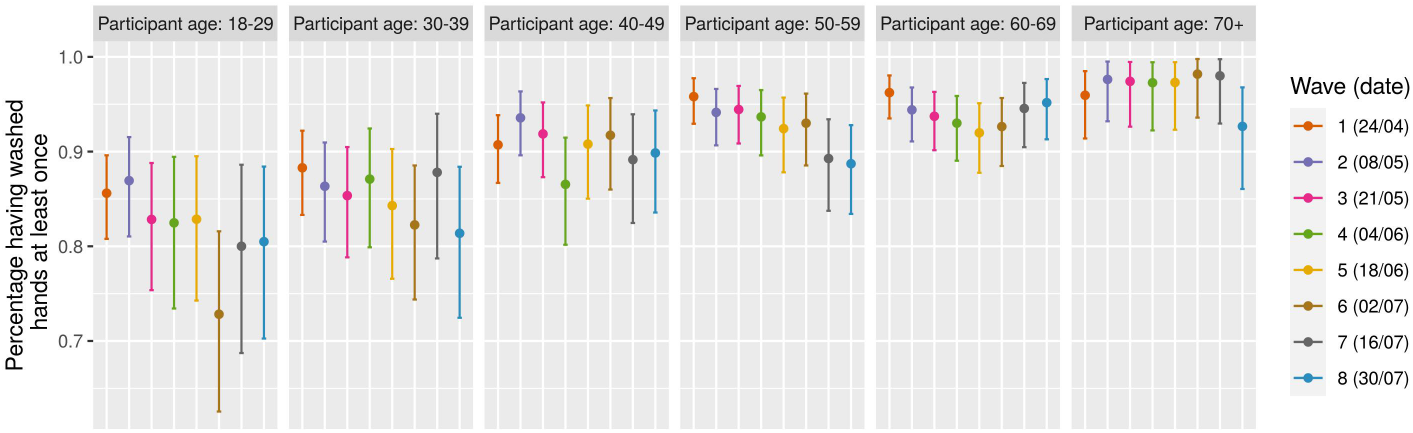
Percentage of participants having washed their hands. Average number of participants having washed their hands at least once in the last three hours, broken down by age. Errorbars mark the 95% CI.

## 4 Discussion

The Belgian government imposed several measures in a sequential manner. On March 13, a directive to close all schools, restaurants, cafes, gyms, and to forbid public gatherings was issued. Further measures followed on March 17th, when more strict limitations on the mobility of people were imposed in addition to closing companies/premises offering non-essential services. On the 4th of May, the Belgian government started to relieve the country lockdown, with a plan of progressive relief for the following weeks. Monitoring how a population complies with social distancing and how it will adjust to the new governmental directions is crucial to predict epidemic scenarios for the months to come.

In this work we report results for the first eight waves of the CoMix survey, aimed at monitoring population compliance and contact patterns in the population. We quantified the impact of social distancing measures on contact patterns, collecting additional information on the use of face masks and compliance to hygiene recommendations.

Although the average number of contacts increased during the different waves of the survey, its value is still significantly lower than pre-pandemic values [11, 12]. During the first two waves of the survey, the reduction in the average number of contacts was around 80% and was quite consistent across all age-classes. This value is in line with what was measured in European countries [6, 10, 7, 8] but smaller than values reported in Shanghai and Wuhan [5] and higher than preliminary results for the US [9]. This reduction in the number of contacts shows a strong location-specific structure. In the lockdown scenario (wave 1), contacts at home account for *∼* 70% of the total number of contacts, with contacts during leisure activities, transportation, and other places accounting for the rest. Therefore, a smaller reduction is measured in the number of contacts at home with respect to all other type of contacts. This is quite reasonable, as social distancing measures impact mostly contact patterns outside households. In the following waves of the survey (waves 3 to 7), the number of contacts presented an increase in the average value. During wave 8, a reduction in the average number of contacts with respect to wave 7 was observed, in line with additional social distancing rules imposed by the government at the end of July [22]. Median values were however quite stable, implying that the distribution of the number of contacts for later waves is more skewed than during lockdown and the first phase of re-opening. Furthermore, the largest increase in the number of contacts is measured for the youngest age class (from 18 years of age to 29 years of age) and in participants from the Flemish region.

The reduction in the number of contacts due to social distancing impacts the reproduction number [13]. With respect to that, we measured the R_0_ ratio considering only adults or imputing contacts of children (i.e. individuals *<*18y). The two approaches lead somehow different results, with higher ratio obtained when imputing children contacts. These results stress that care should be taken when imputing contacts in children based on behavioral changes in adults. However, epidemiological implications in the case of the COVID-19 are difficult to ascertain, as the role of children in the spread of COVID-19 is quite debated [23, 24, 25, 26]. While some studies report a smaller number of confirmed cases in children [27], pointing towards either a reduced susceptibility or a reduced infectivity [5, 27], others report higher infection rates in children with respect to adults [26] and a relevant role in diffusion of children [28]. Without the ability to discriminate between the two cases, a scenario analysis is needed. This would reduce the impact of possible bias induced by contact imputation for children, especially for primary school children. The data shows a slow but constant increase in the absolute reproduction number, which from wave 3 has a median value higher than one. It is worth noting that the largest increase in the average number of contacts is measured among the youngest age class surveyed (18-29 years of age). Taken together, these two results could signal the risk of disease resurgence, especially among the younger individuals. Our result show that physical contacts have not increased after the relaxation of distancing measures in individually reported contacts. Computing the R_0_ ratio only using physical contacts leads to a reproduction number that sets below one for all the waves of the survey. We need to stress, however, that physical contact information is missing for most of the group-reported contacts, that are a considerable percentage of the total number of contacts from wave 3 onward. Therefore, this analysis is performed on a small subset of the total number of contacts.

Participation from wave 1 to wave 2 was quite high (82.8%) and still after 16 weeks of data collection around 50% of the panel participant opted for filling the questionnaire. Establishing a loyal panel of participants is of course crucial for monitoring behavioral changes and population compliance to government directions over time.

We measured an increasing use of face-masks in the participants, starting from 18% during lockdown to more than 75% at the beginning of August. This is confirmed also from a widely participated, non-representative survey [29] that was deployed in Belgium following the lockdown. This increase is likely linked to the increase of contacts outside the households, where wearing a face-mask could be compulsory or perceived as more important to ensure safety during interactions. We did not measure gender-specific effects, although these have been reported in other countries, where face mask wearing was not mandatory [30]. Our work is based on several working assumptions, that may result in limitations of our work. To avoid longer ethical scrutiny and deploy the survey as promptly as possible we decided to exclude children (i.e. individuals *<*18y) from our survey. As we have shown, however, contact imputation for children based on contact reductions of adults leads to inconsistent results, and thus the best option was to limit our considerations to the adult population. Nonetheless, we expect the impact of the younger population to be less crucial for COVID-19 spread. In addition to that, parallel solutions to specifically address children and their contact patterns during school closure terms may be put in place [29]. Our survey asked participants to record contacts retrospectively. Although this could be associated with recalling bias [3, 31], such an effect is marginal when participants have to remember contacts from recent time [32]. This is even more so in a lockdown situation, when the total number of contacts is very low. As a final remark we measured the expected impact of contact patterns on R_0_. It should be stressed, however, that changes in R_0_ could be due to changes in infection transmissibility, which could be affected by several factors, like the use of face mask, increased caution during conversational contacts and possible direct/indirect effect of climate variables on transmission.

In conclusion, we have shown how a representative survey can measure compliance to social distancing and assess its impact on COVID-19 spread. As neither a vaccine nor adequate therapeutic options are available at this time, monitoring the effectiveness of social distancing recommendations is of paramount importance to properly assess the impact of such interventions and avoid further waves of COVID-19.

## Data Availability

The social contact data is available on Zenodo.

https://doi.org/10.5281/zenodo.4035001

## 5 Acknowledgements

This work received funding from the European Research Council (ERC) under the European Union’s Horizon 2020 research and innovation program (grant agreement 682540 TransMID). This project was funded by the European Union’s Horizon 2020 Research and Innovations Programme - project EpiPose (Epidemic intelligence to Minimize COVID-19’s Public Health, Societal and Economical Impact. LW gratefully acknowledge support from the Fonds voor Wetenschappelijk Onderzoek (FWO) (postdoctoral fellowship 1234620N).

We ackwowledge Quentin Leclerc and Alexis Robert for their contribution to the questionnaire translation.

We gratefully acknowledge all the members of the SIMID-COVID19 (in alphabetical order): Steven Abrams, Phlippe Beutels, Joke Bilcke, Pietro Coletti, Jonas Crévecoeur, Chellafe Ensoy-Musoro, Christel Faes, Nicolas Franco, Tapiwa Ganyani, Niel Hens, Lisa Hermans, Sereina A. Herzog, Cécile Kremer, Elise Kuylen, Pieter Libin, Kirsten Maertens, Geert Molenberghs, Yessika Adelwin Natalia, Minh Hanh Nguyen, Benson Ogunjimi, Oana Petrof, Ziv Shkedy, Andrea Torneri, Pierre Van Damme, Joris Vanderlocht, Bieke Vanhoutte, Johan Verbeek, Sarah Vercruysse, Frederik Verelst, James Wambua, Lander Willem.

## 6 Study contribution

Study conceptualisation: PC, JW, NH; Literature research: PC; Data collection: AG,CIJ, KVZ; Data analysis code: PC, AG, LW; Data Analysis: PC, JW, AG; Survey Questionnaire: NH, PB, SV, BV, CJ, KVZ, JE; Manuscript first draft: PC, JW, NH, PB;Coordination: NH. All authors interpreted the findings, contributed to writing the manuscript, and approved the final version for publication.

