## Supporting information for "CoMix: comparing mixing patterns in the Belgian population during and after lockdown"

### ADDITIONAL MATERIAL FOR CoMix: comparing mixing patterns in the Belgian population during and after lockdown

October 23, 2020

[1]Data Science Institute, I-BioStat, UHasselt, Hasselt, Belgium. [2]London School of Hygiene and Tropical Medicine, London, UK [3]Centre for Health Economic Research and Modelling Infectious Diseases, Vaccine & Infectious Disease Institute, University of Antwerp, Antwerp, Belgium. [4]School of Public Health and Community Medicine, The University of New South Wales, Sydney, Australia.

Table 1: **Quotas on age, gender and region.** Quotas on age, gender and region. The target value is compared with the achieved value during wave 1.

| | Age | |
|---|---|---|
| | Achieved | Target |
| 18-24 | 10% | 10% |
| 25-34 | 15% | 16% |
| 35-44 | 16% | 16% |
| 45-54 | 20% | 18% |
| 55-64 | 18% | 16% |
| 65+ | 21% | 23% |
| | Gender | |
| Female | 53% | 49% |
| Male | 47% | 51% |
| | Region | |
| Brussels | 7% | 10% |
| Flanders | 62% | 58% |
| Wallonia | 31% | 32% |

Table 2: **Participation rate.** Number of people invited to each wave and number of people that completed the survey, divide among newly invited and participants from previous wave.

| Wave | New invitations sent | New invitations completed | Participants from previous wave invited | Participants from previous wave completed | Final sample size | Participation (full) | Participation (from previous) |
|---|---|---|---|---|---|---|---|
| 1 | 5978 | 1542 | - | - | 1542 | 25.8% | - |
| 2 | 417 | 94 | 1542 | 1183 | 1277 | 65.2% | 76.7% |
| 3 | 715 | 39 | 1636 | 1103 | 1142 | 48.6% | 67.4% |
| 4 | 0 | 0 | 1675 | 951 | 951 | 56.8% | 56.8% |
| 5 | 0 | 0 | 1675 | 924 | 924 | 55.2% | 55.2% |
| 6 | 0 | 0 | 1675 | 902 | 902 | 53.3% | 53.3% |
| 7 | 0 | 0 | 1675 | 760 | 760 | 45.4% | 45.4% |
| 8 | 0 | 0 | 1675 | 833 | 833 | 49.7% | 49.7% |

### 1 Supporting information

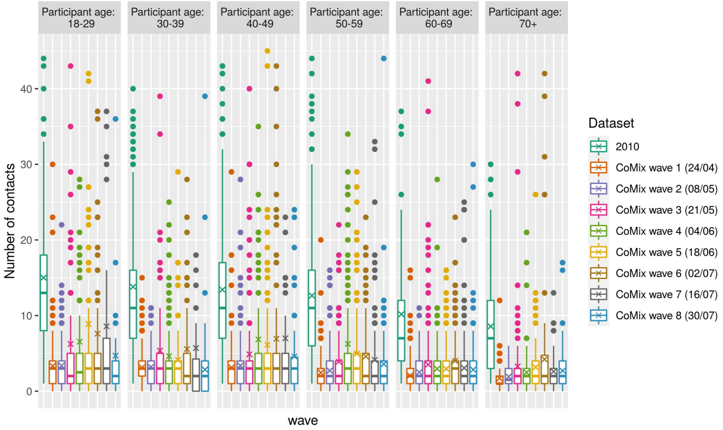


Figure 1: **Number of contacts.** Boxplot of the number of contacts. The boxplot marks the interquartile range (IQR), The horizontal line marks the median value and the 'x' symbol marks the average. Whiskers extend up to 1.5 times IQR and outliers shown as point.

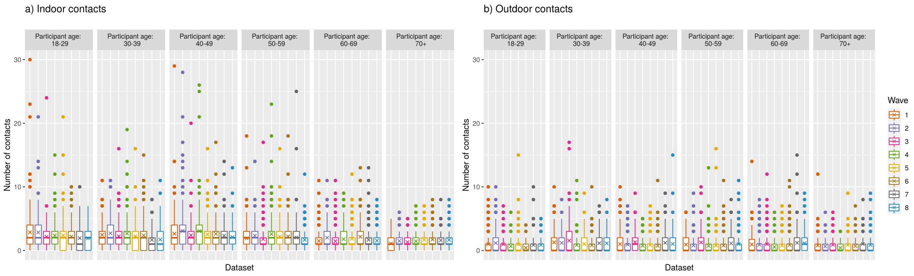


Figure 2: **Indoor/outdoor contacts (a)**: Indoor contacts.**(b)**: Outdoor contacts. In both panels, the boxplot shows the interquartile range (IQR), The horizontal line marks the median value and the 'x' symbol marks the average. Whiskers extend up to 1.5 times IQR and outliers are shown as point.

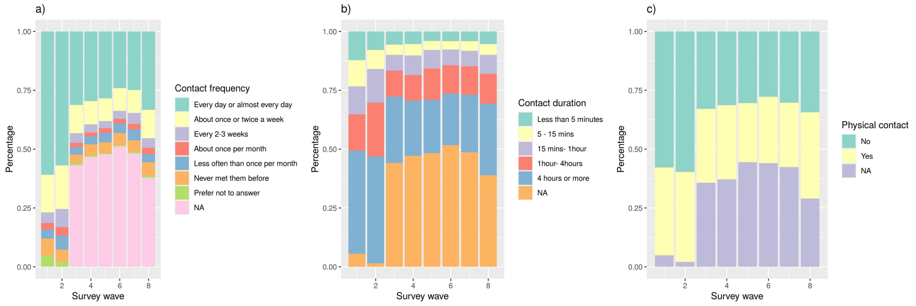


Figure 3: **Contact features. (a):** Contact frequency.**(b):** Contact duration.**(c):** Physical contact. From wave 3 onward, a large fraction of contacts are reported in group, missing information on contact frequency, duration and physical contact.

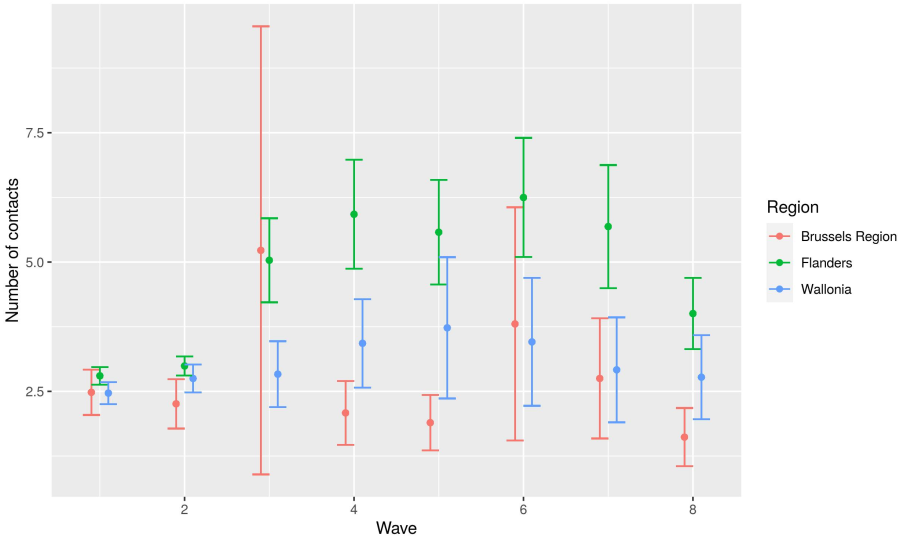


Figure 4: **Average number of contacts broken down by geographical area.** Mean number of contacts broken down by geographical area. Errorbars mark the 95% CI.

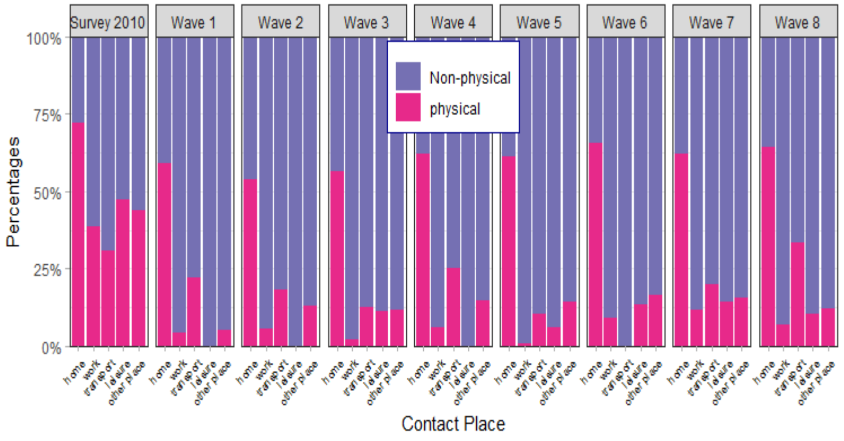


Figure 5: Proportion of contacts that involved physical contact by contact place for the 2010 survey in Flanders and the 2020 CoMix surveys (only individually reported contacts)

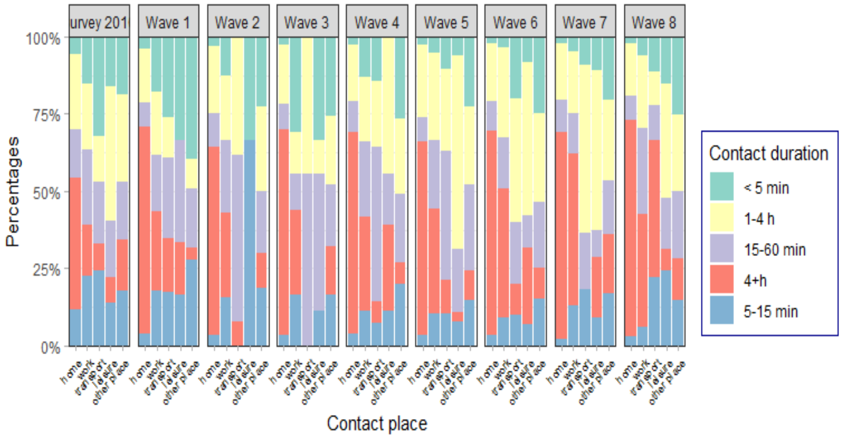


Figure 6: The distribution of all contacts by contact duration and contact place for the 2010 survey in Flanders and the 2020 CoMix surveys (only individually reported contacts)

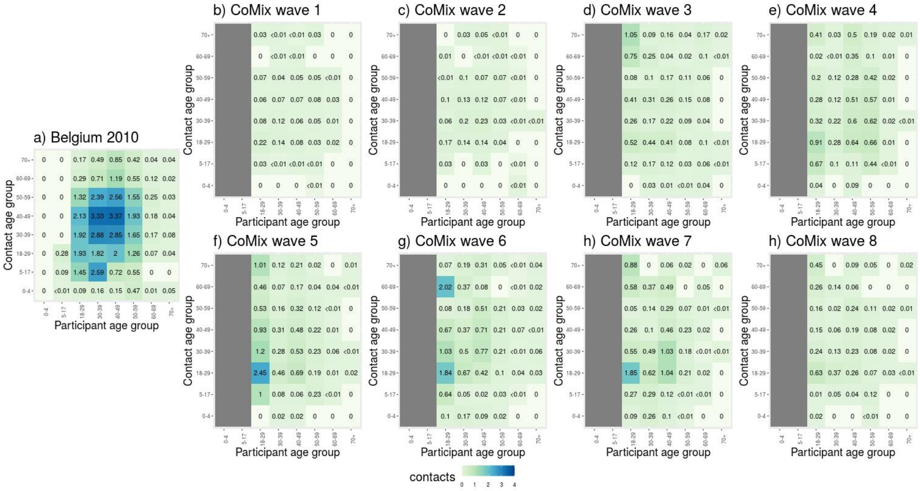


Figure 7: **Social contacts at work. (a):** Average number of daily contacts at work for the 2010 survey. **(b-i)** Average number of reported contacts at work for the 8 waves of the CoMix survey.

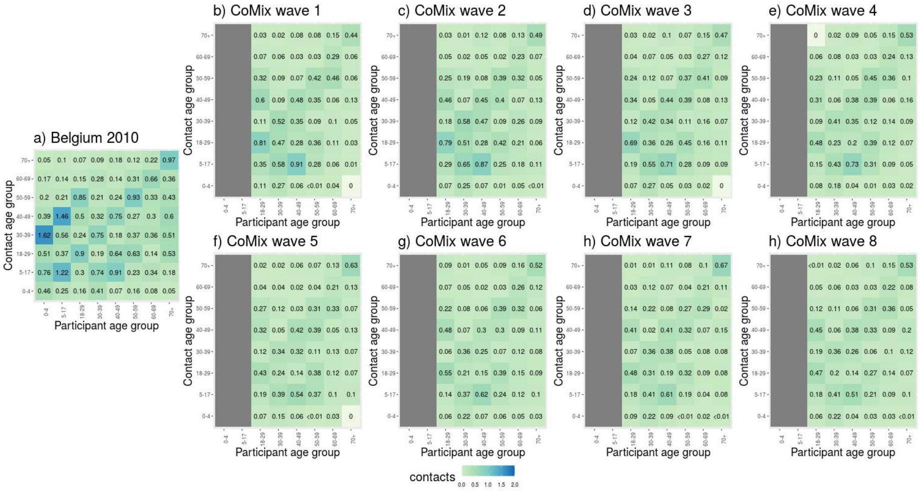


Figure 8: **Social contacts at home. (a):** Average number of daily contacts at home for the 2010 survey. **(b-i)** Average number of reported contacts at home for the 8 waves of the CoMix survey.

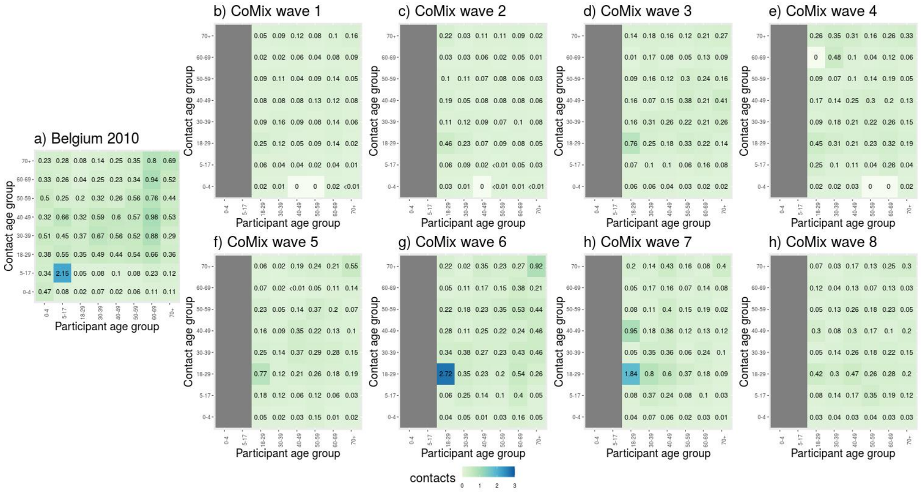

Figure 9: **Social contact “other” locations. (a):** Average number of daily contacts made at “other” locations for the 2010 survey. **(b-i)** Average number of reported contacts made at “other” locations for the 8 waves of the CoMix survey.

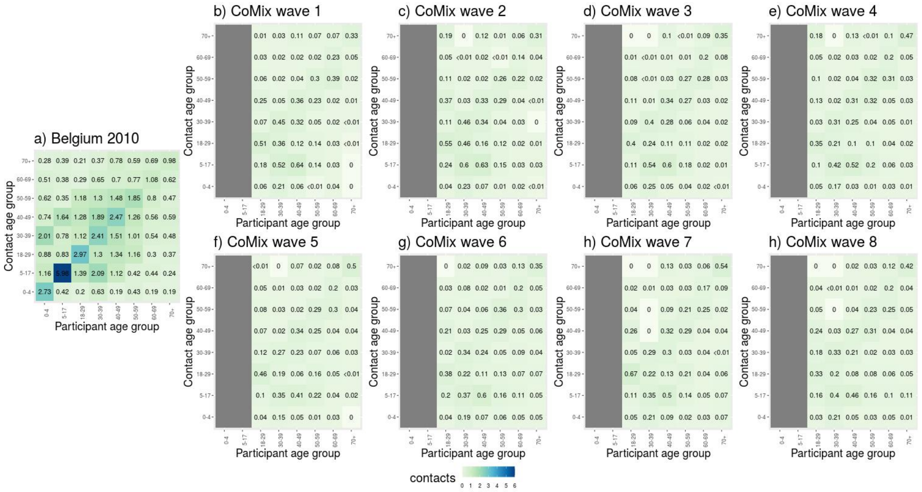

Figure 10: **Physical contacts. (a):** Average number of daily physical contacts for the 2010 survey. **(b-i)** Average number of reported physical contacts for the 8 waves of the CoMix survey.

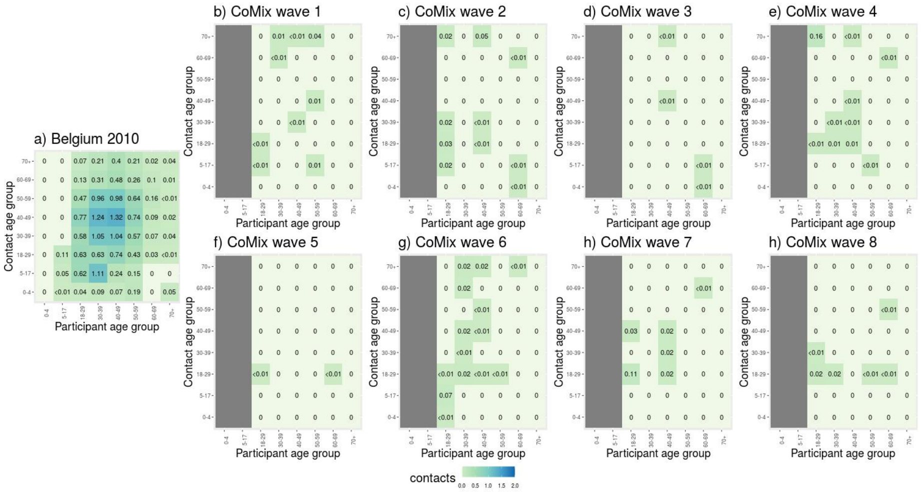


Figure 11: **Physical contacts at work. (a):** Average number of daily physical contacts at work for the 2010 survey. **(b-i)** Average number of reported physical contacts at work for the 8 waves of the CoMix survey.

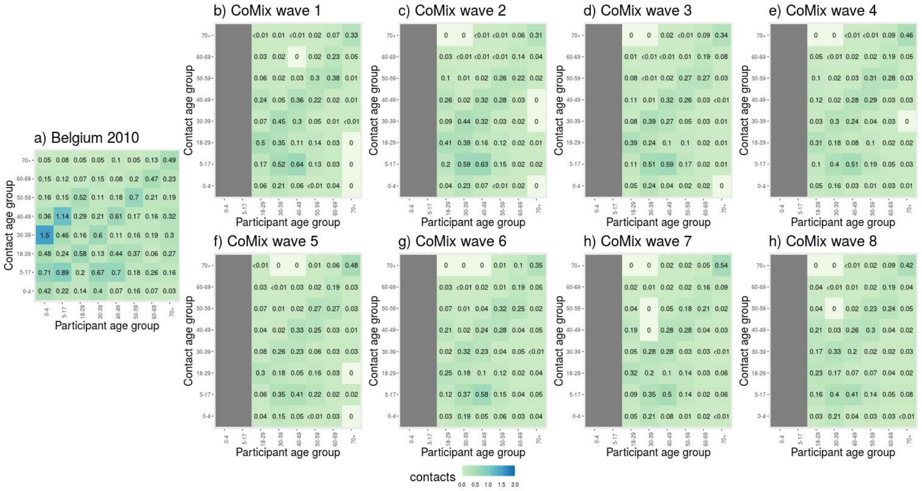


Figure 12: **Physical contacts at home. (a):** Average number of daily physical contacts at home for the 2010 survey. **(b-i)** Average number of reported physical contacts at home for the 8 waves of the CoMix survey.

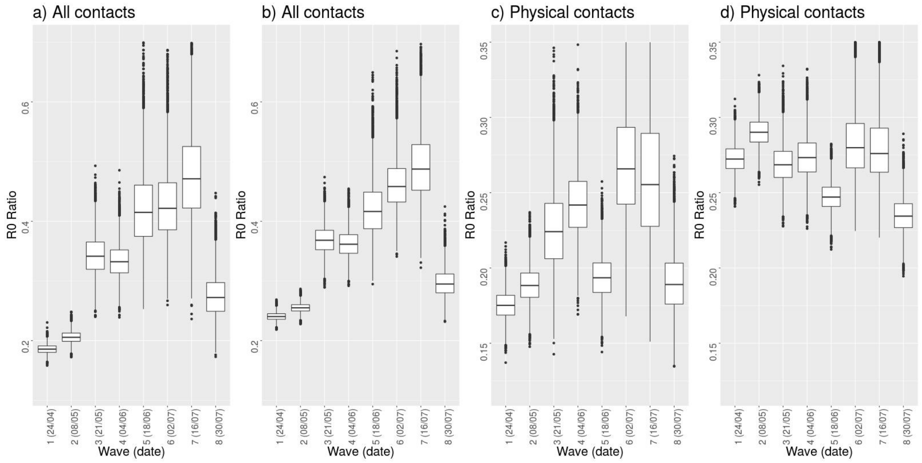


Figure 13: **$R_0$ ratio. (a-b):** $R_0$ ratio considering all contacts, without imputation (a) and with imputation (b).**(c-d):** $R_0$ ratio considering only physical contacts, without imputation (c) and with imputation (d). The ratio of $R_0$ is computed with respect to the 2010 survey data. In both panels, the boxplot shows the interquartile range (IQR), The horizontal line marks the median value and the 'x' symbol marks the average. Whiskers extend up to 1.5 times IQR and outliers shown as point.

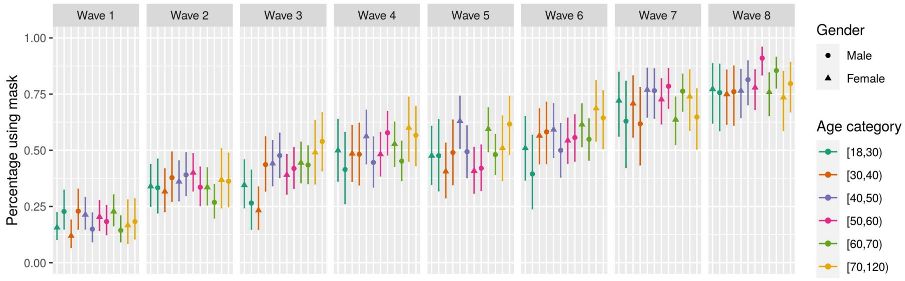


Figure 14: **Face mask use broken down by age and gender.** Percentage of participants wearing mask broken down by age and gender. Errorbars mark the 95% CI.

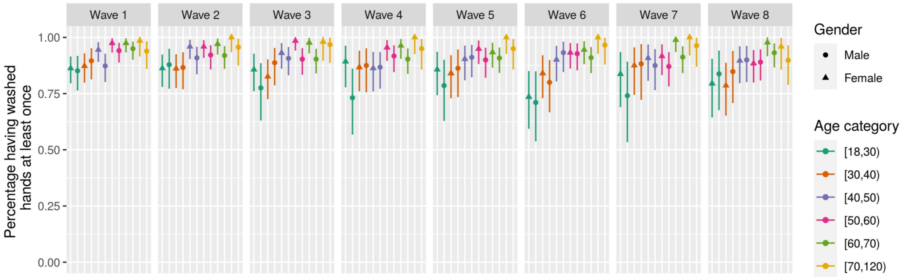


Figure 15: **Percentage of participants having washed their hands broken down by age and gender.** Percentage of participants having washed their hands broken down by age and gender. Errorbars mark the 95% CI.

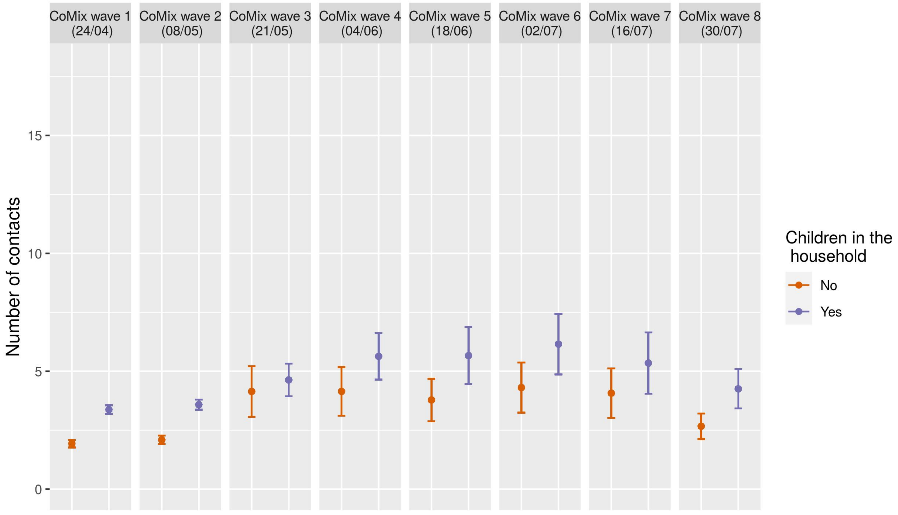


Figure 16: **Average number of contacts according to household structure.** Mean number of contacts for households including and not including a children. Errorbars mark the 95% CI.
